# Patient-Level Stroke Outcomes Following Linkage of a Statewide Stroke Registry to Administrative Claims Data

**DOI:** 10.1101/2025.11.03.25339450

**Authors:** Raed Hailat, Michael P Thompson, Adrienne Nickles, J. Adam Oostema, Gustavo de los Campos, Mathew J Reeves

## Abstract

**Background:** Collection of patient-level outcomes data following hospital discharge is challenging for stroke registries. Data linkage to administrative claims data is a potential solution to obtain outcomes data. We aimed to generate data on 30-day, 90-day and 1-year outcome events following hospitalization for stroke using linked data in Michigan.

**Methods:** We probabilistically linked clinical data from a 5-year cohort (2016-2020) of all index acute stroke discharges (ICD-10 I61-I63) from 31 hospitals participating in Michigan’s Acute Stroke program (MiSP) to a representative statewide multi-payer claims database. We used the linked data to generate data on 30-day, 90-day, and 1-year event rates including hospital readmissions, stroke recurrence, post-acute care services (i.e., facility-based rehabilitation and home health), and out-patient visits. Mortality data was only available for Medicare fee-for-service beneficiaries. Outcomes were stratified by age, race, stroke type, and stroke severity.

**Results:** Of the 46,330 MiSP stroke discharges, 23,918 (51.6%) were linked to the claims database. Readmission and stroke recurrence rates were 14.1% and 3.3%, respectively, at 30 days, increasing to 42.2% and 8.3% at one year. By 30 days about a quarter of subjects had used facility-based rehab and another quarter had used home health; home health utilization increased to 44.7% by one year. At all time points Black patients had significantly higher readmission rates compared to whites, but higher stroke recurrence rates were only observed at the 1-year mark. At 30 days, utilization of post-acute care services did not differ by race, but utilization rates were significantly higher in Blacks at 90 days and one year. In contrast utilization of outpatient services was significantly higher among White patients at all time points.

**Conclusions:** Linkage between acute stroke registry and claims data provides an important source of surveillance data for stroke outcomes up to 1-year post discharge. This data allows for real-time monitoring of healthcare outcomes and potentially leads to interventions to improve stroke care.

## Introduction

The large volume of data collected in the national Get With The Guideline-Stroke (GWTG-S) program–now exceeding 9 million stroke discharges from more than 2,600 hospitals has allowed for the detailed examination of trends in quality of stroke care and outcomes up to the point of hospital discharge.^1, 2^ Yet, for stroke patients the majority of functional recovery and return to community participation occurs after hospital discharge.^3^ Collection of outcomes data following hospital discharge has been difficult for large stroke registries to sustain, but a potentially feasible and sustainable way to track patient outcomes is through data linkage to administrative databases that can provide information on patient outcomes including mortality, readmissions, recurrence and post-acute care utilization.^4-6^ Data linkage can serve an important surveillance function by providing real-time data on cardiovascular disease that can be used to track patient outcomes and improve population health.^7^

Therefore, using linked data from a 5-year cohort of acute stroke discharges from a representative stroke registry in Michigan, we aimed to generate descriptive data on 30-day, 90-day and 1-year outcome event rates and stratify the data by age, race, stroke type, and stroke severity.

## Methods

This report adheres to Strengthening the Reporting of Observational Studies in Epidemiology (STROBE) guidelines.^8^ The study was based on 5-year cohort of prospectively collected data of index acute ischemic and hemorrhagic stroke discharges (ICD-10 I61-I63) collected between January 2016 and December 2020 by 31 hospitals participating in the Michigan Stroke Program (MiSP). The MiSP data was probabilistically linked to a representative statewide claims database (Michigan Value Collaborative [MVC]) that included Medicare fee-for service (FFS) and private insurer data, using date of birth, sex, admission date, discharge date, and hospital ID. Both MiSP and MVC datasets were deidentified. Clinical data including demographics (age, race, sex), stroke type (ischemic and hemorrhagic), discharge disposition, and stroke severity (National Institute of Health Stroke Scale [NIHSS]) were based on MiSP data. The MVC dataset included all post-acute health services claims submitted to the insurance provider during the 12-month period following hospital discharge for the index event. These services include services (in-patient rehabilitation (IRF), skilled nursing facility (SNF), home health care (HHC), outpatient medical visits (OP), and hospital readmissions. Mortality data were only available for FFS beneficiaries. More information on study databases is available in the supplement.The index stroke events were defined as the patient’s first-stroke discharge during the 5-year study period. We excluded patients that died during hospitalization or were discharged to hospice care or against medical advice (Figure S1 Supplement).The claims data were used to calculate 30-day, 90-day and 1-year event rates of all-cause hospital readmissions-defined as any subsequent hospitalization within one year of the discharge date of the index event, stroke recurrence-defined as any subsequent stroke-related hospital discharge occurring within one year of the index stroke, use of post-acute care services (i.e., IRF, SNF, and HHC), and use of OP visits-defined as any outpatient claim for primary or specialty care. Home time-defined as post discharge time spent alive and out of an inpatient care setting (i.e., hospital, IRF, SNF, or Long term acute care hospital), was only calculated for FFS beneficiaries since mortality data were available for this group.^9^ One year Kaplan-Meier curves for mortality, all-cause readmission, and stroke recurrence were generated. We stratified outcomes according to age, race, stroke type and severity. Data analysis was done using SAS software v9.4 (Cary, NC).

## Results

Data linkage was conducted between the 46,330 MiSP and 30,685 MVC acute stroke discharges and resulted in 19,382 matched index strokes. The final linked cohort were predominantly older(>65 years, 78.5%), white (79.7%), female (52.2%), and insured by Medicare FFS (62.9%) (Table 1). The majority of index hospitalizations were ischemic stroke (87.3%), with a predominance of minor stroke (NIHSS <5, 56.4%). About half of index stroke hospitalizations were discharged home, 21.4% were discharged to IRF, and 24.9% to SNF. A total of 14.1%, 24.9%, and 42.2% of the linked population were readmitted at least once within 30 days, 90 days, and one year post discharge, respectively (Table 2 and Figure 1A). Only 3.3% of the linked population had a recurrent stroke event (defined as a hospital readmission with a primary discharge diagnosis of stroke) within 30 days; this increased to 5.1% at 90 days and to 8.3% at one year (Table 2 and Figure 1B).

**Table 1:** Descriptive characteristics of a 5-year cohort of linked index stroke events (N=19,382).

| Variable | Value | Distribution N (%) |
| --- | --- | --- |
| Age category | <65 | 4,167 (21.5) |
|  | 65-74 | 5,672 (29.2) |
|  | 75-84 | 5,589 (28.8) |
|  | >=85 | 3,954 (20.4) |
| Race | White | 15,457 (79.7) |
|  | Black | 2,833 (14.6) |
|  | Other* | 255 (1.3) |
|  | Not documented | 837 (4.3) |
| Latino ethnicity | No/Unable to determine | 18,665 (96.3) |
|  | Yes | 717 (3.7) |
| Sex | Male | 9,265 (47.8) |
|  | Female | 10,117 (52.2) |
| Insurance | Private (Preferred Provider Organization (PPO) and Health Maintenance Organization (HMO)) | 3,140 (16.2) |
|  | Medicare Advantage (MA) | 4,051 (20.9) |
|  | Medicare Fee For Service (FFS) | 12,191 (62.9) |
| Stroke type | Ischemic | 16,921 (87.3) |
|  | Hemorrhagic | 2,461 (12.7) |
| Admission<br>NIHSS | 0 | 2,985 (15.4) |
|  | 1-4 | 7,937 (41.0) |
|  | 5-15 | 4,955 (25.6) |
|  | 16-20 | 830 (4.3) |
|  | >20 | 733 (3.8) |
|  | Not documented | 1,942 (10.0) |
| Discharge<br>disposition | Home | 9,710 (50.1) |
|  | Skilled Nursing Facility (SNF) | 4,821 (24.9) |
|  | Inpatient Rehabilitation Facility (IRF) | 4,148 (21.4) |
|  | Long Term Care Hospital (LTCH) | 122 (0.6) |
|  | Not determined | 581 (3.0) |

\*Includes American Indian or Alaska Native, Asian, and Native Hawaiian or Other Pacific Islander.

**Table 2:**
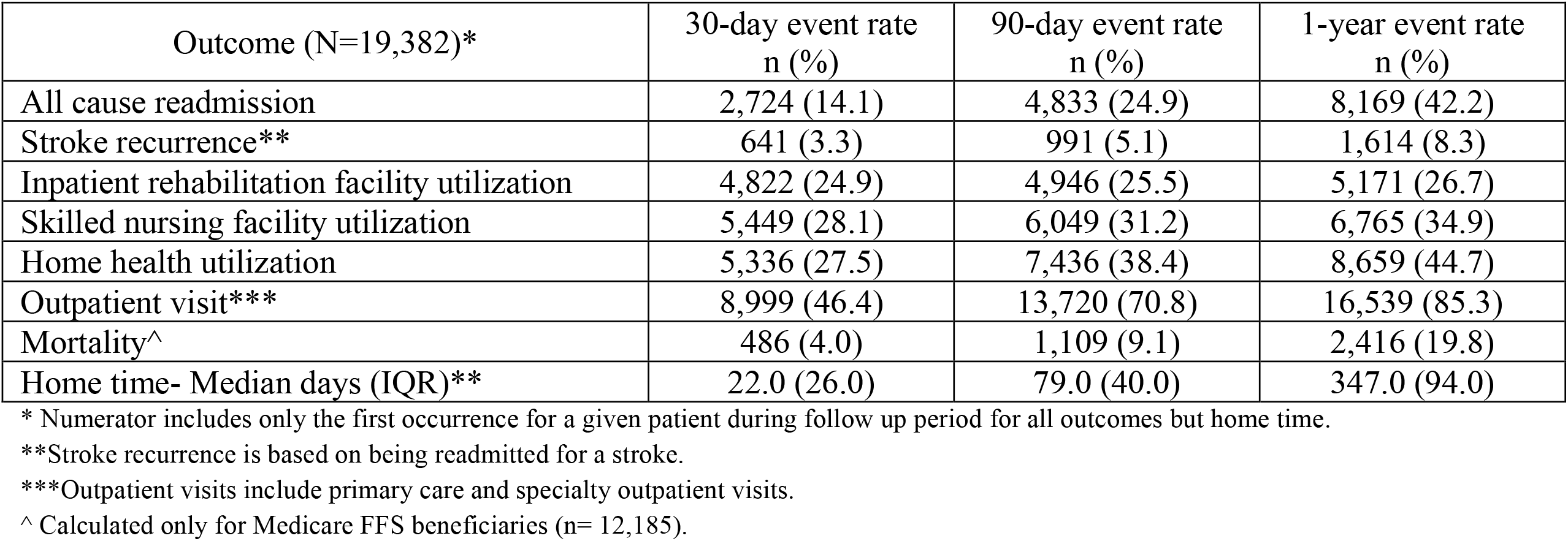
Thirty-day, 90-day, and 1-year post stroke discharge outcome event rates among a 5-year cohort of 19,382 linked stroke events.

**Figure 1.**
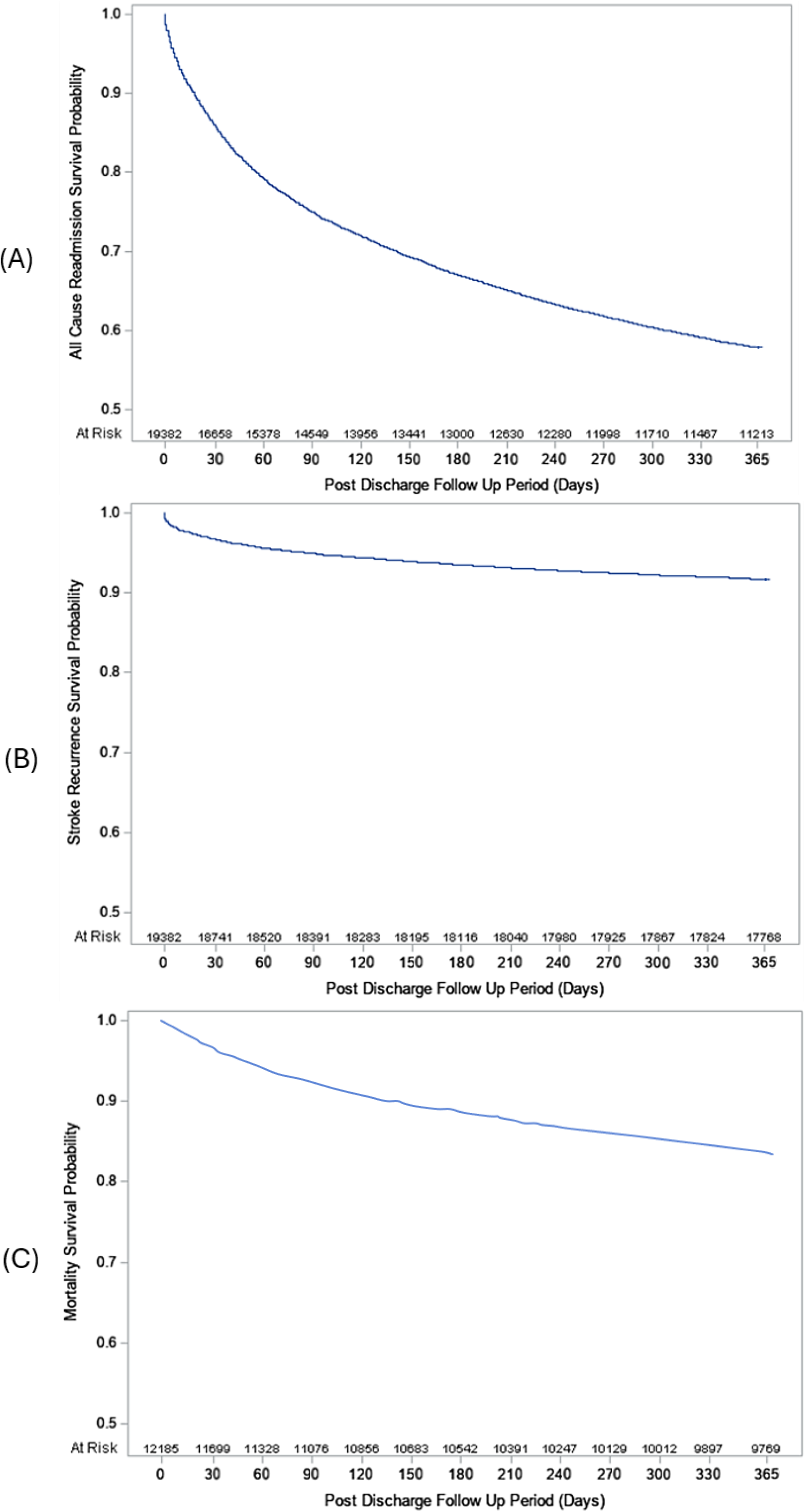
Kaplan-Meier curves of all cause readmission (1A), stroke recurrence (1B), and mortality (1C).

In terms of use of post-acute services, 24.9%, 28.1%, 27.5% of the linked cohort utilized IRF, SNF, and HHC within 30 days of hospital discharge, respectively (Table 2). While IRF and SNF utilization rates increased only slightly at 90 days (25.5% and 31.2%, respectively), HHC utilization increased substantially to 38.4% at 90 days and 44.7% by one year (Table 2). By 30 days, 90 days, and one year of follow-up, 46.4%, 70.8%, and 85.3% of the cohort had at least one OP visit, respectively.

For the 12,185 Medicare FFS linked index events, mortality rates were 4.0%, 9.1%, and 19.8% at 30 days, 90 days, and one year post discharge, respectively (Table 2 and Figure 1C). Median home time was 22, 79, and 347 days at 30 days, 90 days, and one year, respectively (Table 2). Compared to the <65 years old group (n= 4,167), the ≥65 years old group (n= 15,215) all cause readmission rates were higher at 90 days and one year of follow up (Table S1), however, stroke recurrence among the ≥65 years old group was consistently lower compared to the <65 years old group. SNF utilization was higher among the ≥65 years old group over the entire one year followup period, but IRF utilization did not differ; OP services were utilized at a higher rate up to 90 days post discharge compared to the <65-year-old group.

Compared to the Whites (n= 15,457), the Blacks (n= 2,833) had higher all-cause readmission rates over the entire follow up period, whereas stroke recurrence rates in Blacks were only significantly higher at one year (Table S2). Blacks also had higher IRF, SNF and HHC utilization rates over the one year of follow up, but they utilized OP services less often than the Whites.

Although mortality rates among the Medicare FFS beneficiaries were similar across racial groups, blacks had a consistent lower home time over one year of follow up reflecting their higher rates of readmission and use of facility based post-acute care services (IRF and SNF). Compared to the hemorrhagic stroke patients, ischemic stroke patients had lower all-cause readmission and stroke recurrence rates over the entire one year follow up (Table S3). IRF and SNF utilization rates were also lower among ischemic stroke patients compared to hemorrhagic stroke patients. Among FFS beneficiaries, ischemic stroke patients had lower mortality rates and greater home time compared to hemorrhagic stroke patients over one year.

Patients with severe strokes (NIHSS >15) had the highest all-cause readmission rates but stroke recurrence did not differ by severity (Table S4). Utilization of health care services was strongly impacted by stroke severity; IRF and HHC utilization rates were the highest among patients with moderate stroke (NIHSS 5-15), while SNF utilization was highest in patients with severe stroke patient group, and OP utilization was highest among mild stroke (NIHS < 5). Among FFS beneficiaries, patients with severe stroke had the highest mortality rates and lowest home time over one year.

## Discussion

We generated descriptive data on 30-day, 90-day and 1-year outcome event rates using linked data from a 5-year cohort of acute stroke discharges from Michigan. Our data fulfills the American Heart Associations call for real time collection and analysis of cardiovascular disease surveillance data which is important to identify at-risk groups, and drive advancements in prevention and treatment strategies, including precision health and policy development.^7^The overall stroke outcome event rates generated in this analysis were generally similar to those previously published. Our 30-day and 1-year readmission rates were very similar to those reported by a recent meta-analysis that found pooled 30-day and 1-year all-cause stroke readmission rates of 17.4% and 42.5%, respectively.^10^ Our 30-day, 90-day, and 1-year stroke recurrence rates were also similar to a report that examined 2017 Medicare FFS cases with ischemic stroke that found 30-day, 90-day, and 1-year recurrence rates of 2.4%, 4.0%, and 7.6%, respectively.^11^

With respect to health care utilization post hospital discharge, our 30-day utilization rates for IRF were similar but SNF and HHC rates were above what a nationally representative GWTG-S population study conducted between 2003 and 2011 reported on.^12^ However, the GWTG-S study collected their data at the point of discharge, whereas our data was based on actual claims accrued during the 30 days post discharge period which explains the higher 30-day rates of SNF and HHC utilization. We observed increasing rates of SNF and HHC utilization from 30 days to 90 days which points to the fact that many patients initially discharged without these services end up using them. Regarding OP visits, our rates are similar to utilization rates reported by a nationally representative claims database of commercially insured Americans from 2009 to 2015, where 70.8% of acute stroke patients had a primary care visit within 90 days post discharge.^13^

The value of obtaining patient outcome data from data linkage has been illustrated by previous studies that have linked the GWTG-S registry to Medicare fee-for-service (FFS) data.^14-22^ However, these reports do not include stroke patients younger than 65 years or Medicare Advantage beneficiaries which are a rapidly growing population.^5, 23-25^ Data on post discharge OP medical visits utilization is also very important to track adherence to OP treatment and secondary prevention efforts.

Of the remaining outcomes examined, our 30-day, 90-day, and 1-year mortality rates were similar to rates of prior FFS population with minor stroke, where 30-day, 90-day and 1-year all-cause mortality were reported as 3.7%, 7.6%, and 17.2%, respectively.^20^ The calculated median 90-day and 1-year median home time estimates of 79 and 347 days were similar to prior data report that also used Medicare FFS population (90-day: 79 days; 1-year: 349 days).^17^ Home time can be used to track stroke recovery post discharge as it is recognized as a valid proxy measure of functional recovery in prior studies.^17, 26-30^

The ability to stratify the outcomes data by race and stroke severity shed light at the importance of data linkage in enabling these stratifications which are usually not able to be generated from claims data alone. It also emphasizes the important differences in event rates and use of post discharge services across subgroups which is relevant to developing patient specific post discharge follow up plans.^31, 32^ This study found important difference by age and race. We found as expected that older age groups had higher readmission rates, but it was unexpected to find lower stroke recurrence rates in older subjects. Prior studies of stroke recurrence in the young versus older patients are relatively unusual; the South London Stroke registry reported no difference in stroke recurrence rates between <65 and ≥65 years old patients.^33^ Our one year stroke recurrence rate in the younger (<65) age group is similar to that of a meta-analysis of 31 studies that reported a recurrence rate of 7.3%.^34^

We also found important differences by race, Black patients had higher hospital readmission rates and use of IRF, SNF, and HHC services compared to White patients, however they were more likely to get readmitted with a recurrent stroke only at one year. This might reflect poorer adherence to secondary prevention treatments, especially given that we found that they utilized OP services less often than white patients. Previous studies have also found lower medication adherence following stroke in black patients which could also contribute to higher recurrence rates.^35, 36^

Our study has some limitations to consider. The data is based on linked data from 31 stroke accredited hospitals in Michigan, and so findings might not extrapolate to non-stroke certified hospitals.^37^ The data linkage process may have produced biased estimates because we were not able to link to patients in the registry that had insurance coverage outside of the claims data coverage (i.e., MA and commercial plans from other private insurers and Medicaid), however the generated stroke outcomes were generally similar to previously published rates in the literature. In summary, we demonstrated that linkage between acute stroke registry and claims data allowed for reporting of several key stroke outcome metrics up to 1-year post discharge. This data provides important insights into the use of medical services and patient outcomes that can be incorporated into population-based surveillance data and further studied by healthcare professionals, systems, and policy makers to develop interventions, and evaluations that can potentially lead to improvements in stroke care.

## Data Availability

The data used in this study were obtained under multiple data use agreements that prohibits public sharing. Access to these data can be requested directly from The University of Michigan- Michigan Value Collaborative and Michigan Department of Health and Human Services- Michigan Stroke Registry following their established procedures.

## Acknowledgments

None

## Sources of Funding

Supported by American Heart Association grant #909423/Dr. Raed Hailat/2022

## Disclosures

Support for the Michigan Value Collaborative is provided by Blue Cross Blue Shield of Michigan as part of the BCBSM Value Partnerships program; however, the opinions, beliefs and viewpoints expressed by the author do not necessarily reflect those of BCBSM or any of its employees.

## Notes

### Competing Interest Statement

The authors have declared no competing interest.

### Clinical Trial

NA

### Author Declarations

IRB approvals were obtained from Michigan Department of Health and Human Services, Michigan State University, and University of Michigan.

